# Uptake of SARS-CoV-2 workplace testing programs, March 2020 to March 2021

**DOI:** 10.1101/2021.06.29.21259730

**Authors:** Nathan Duarte, Sean D’Mello, Natalie A Duarte, Simona Rocco, Jordan Van Wyk, Abhinav Arun Pillai, Michael Liu, Tyler Williamson, Rahul K Arora

## Abstract

**Objective:** To track uptake of workplace SARS-CoV-2 testing programs using publicly-available data (e.g., press releases), supplementing findings from employer surveys.

**Methods:** We tracked testing programs reported by 1,159 Canadian and 1,081 international employers across sectors from March 1, 2020 to March 31, 2021. We analyzed trends in uptake of testing programs, including over time and by workplace setting.

**Results:** 9.5% (n=110) of Canadian employers and 24.6% (n=266) of international employers tracked reported testing. The prevalence of reported testing programs was less than 20% in some settings associated with high risk of transmission including retail and customer-facing environments, and indoor and mixed blue collar workplaces.

**Conclusions:** Publicly-available data suggest that fewer employers are testing than indicated by surveys. Workplace safety in high-risk workplaces could be further improved by implementing testing strategies that deploy both screening and diagnostic tests.

## INTRODUCTION

Over the course of the COVID-19 pandemic, workplace outbreaks of SARS-CoV-2 amplified community transmission, and widespread business shutdowns damaged economies^1,2^. Across sectors, employers were tasked with creating safe work environments in the face of the changing pandemic and an evolving understanding of SARS-CoV-2 transmission, testing, and countermeasures.

Modeling^3^ and pilot studies^4,5^ have shown that SARS-CoV-2 diagnostic testing programs are effective and feasible tools for limiting workplace transmission. Data on whether and how employers are testing their employees could provide important context for occupational health experts and public health officials as they formulate guidance that shapes employer precautions. However, there is limited data of this sort available. One recent study surveyed 1,339 employers and found that two in three are testing employees^6^, but with a response rate of less than 5%, this result is likely subject to substantial non-response bias: employers carrying out testing programs are invested in workplace safety and may have been more likely to respond.

While the increasing pace of vaccination provides hope for a return to normalcy, continued vigilance by employers is warranted, particularly in high-risk workplaces with low vaccine uptake^7^ and also due to emerging variants of concern^8^. As well, studying the workplace response to the COVID-19 pandemic can help inform improvements in the occupational health response to future public health crises.

We used publicly-available data (e.g., press releases, news articles) to track 2,240 employers for reported workplace testing programs, particularly focusing on Canadian employers (n=1,159) while also monitoring international employers (n=1,081). We aimed to (1) understand what type of testing has been reported, (2) assess the evolution of testing over time, and (3) investigate how testing varied by type of workplace.

## METHODS

### Employer list

Scraping publicly-available data^9^ (e.g., press releases, news articles) is well suited for monitoring testing programs implemented by large employers: these businesses employ disproportionately larger segments of the workforce and are the most likely to publicly report testing programs. To generate a list of employers to track, we drew from stock exchanges (e.g., Toronto Stock Exchange, S&P 500, Brazil’s B3), curated lists that rank employers by size (e.g., Financial Post 500, Fortune Global 2000), and expert recommendations. We then stratified companies from these sources between Canadian and international employers, and selected the largest companies by market capitalization and sector. Our final list included 1,159 Canadian employers and 1,081 international employers, providing granular information about the private sector response within one country and a broad global picture from the largest international employers.

### Stratifying employers by workplace type

Testing programs are most helpful in certain working conditions^3^. For example, settings where employees work indoors without distancing for extended periods of time may particularly benefit from testing given the airborne nature of SARS-CoV-2 transmission^10^. Traditional occupational classification systems (e.g., NAICS) are not designed to capture these differences in workplace risk of SARS-CoV-2 transmission; because of this, we developed an internal system to group employers with similar workplace risk profiles. For example, while indoor blue collar workplaces (e.g., factory floors) and mixed (indoor and outdoor) blue collar workplaces (e.g., construction sites) can both involve work where distancing or use of personal protective equipment is difficult, transmission is more likely in indoor-only environments^10^. Supplementary Table 1 outlines the full set of these categories.

We confirmed our classification system’s reliability by randomly sampling ∼5% (n=112) of employers from the shortlist and having two reviewers with experience working with occupational classification systems (ND and SD) independently classify each employer. Cohen’s kappa statistic, a common measure of interobserver agreement^11^, was 0.93, suggesting “almost perfect” agreement and that the process of assigning employers to a workplace type was repeatable. See Supplementary Table 2 for a complete breakdown of companies tracked by country and workplace type.

### Tracking companies

In order to capture publicly-available data about testing programs instituted by each tracked employer, we used Google’s Search API to query the most relevant online documents (e.g., press releases, news articles, company web pages) for each company. We queried results on an ongoing basis from March 1, 2020 to March 31, 2021. We used the following search phrases (entered without the use of quotation marks): “COMPANY_NAME performing antibody tests on employers” and “COMPANY_NAME performing COVID-19 tests on employers”. Because it would be infeasible to screen the entirety of Google’s repository of search results, we leveraged the fact that Google ranks results based on relevance^12^; we saved the five most relevant results for each phrase, compiling a total of ten search results per company per query.

If a search result indicated that a tracked company had either begun testing employees or had updated their workplace testing program (e.g., to change the type or frequency of testing, or to end testing), we recorded this information in our database. We extracted fields including the date of the report, testing modality (antigen, nucleic acid, or antibody), and the monitoring speed (lab-based versus point-of-care). To ensure that the extracted records were accurate, each entry was reviewed independently in duplicate by a second reviewer.

## RESULTS

As of March 31, 2021, 9.5% (n=110) of Canadian employers and 24.6% (n=266) of international employers had reported initiatives to test employees for current or past infection with SARS-CoV-2. Of these employers, 250 (66.5%) specified the testing modality: 174 (69.6%) used diagnostic testing (64% nucleic acid, 23% antigen, 13% both); 34 (13.6%) used antibody tests only; and 42 (16.8%) used both diagnostic and antibody tests. 167 (44.4%) employers specified whether they were using point-of-care or lab-based diagnostic tests: 49 (29.3%) used only point-of-care tests, 83 (49.7%) used only lab-based tests, and 35 (21.0%) used both.

To understand how the pandemic’s trajectory may have prompted employers to begin testing, we compared the number of new testing programs reported and the number of new confirmed cases. We focused on Canada (see Figure 1) due to the granularity with which we tracked its private sector. It was evident that each pandemic wave spurred a “wave” of adoption of testing programs. The first round of testing adoption was drawn out over multiple months, even beyond the subsiding of the first surge in cases, while the second had a much faster ramp up. Across all employers we tracked, the median date on which antibody testing was reported was June 18, 2020 [IQR: May 6 – August 14, 2020], which was before the median date for nucleic acid testing of July 29, 2020 [IQR: May 28 – October 6, 2020]. Antigen tests were adopted most recently, with a median date of December 18, 2020 [IQR: October 11, 2020 – January 23, 2021].

**Figure 1.**
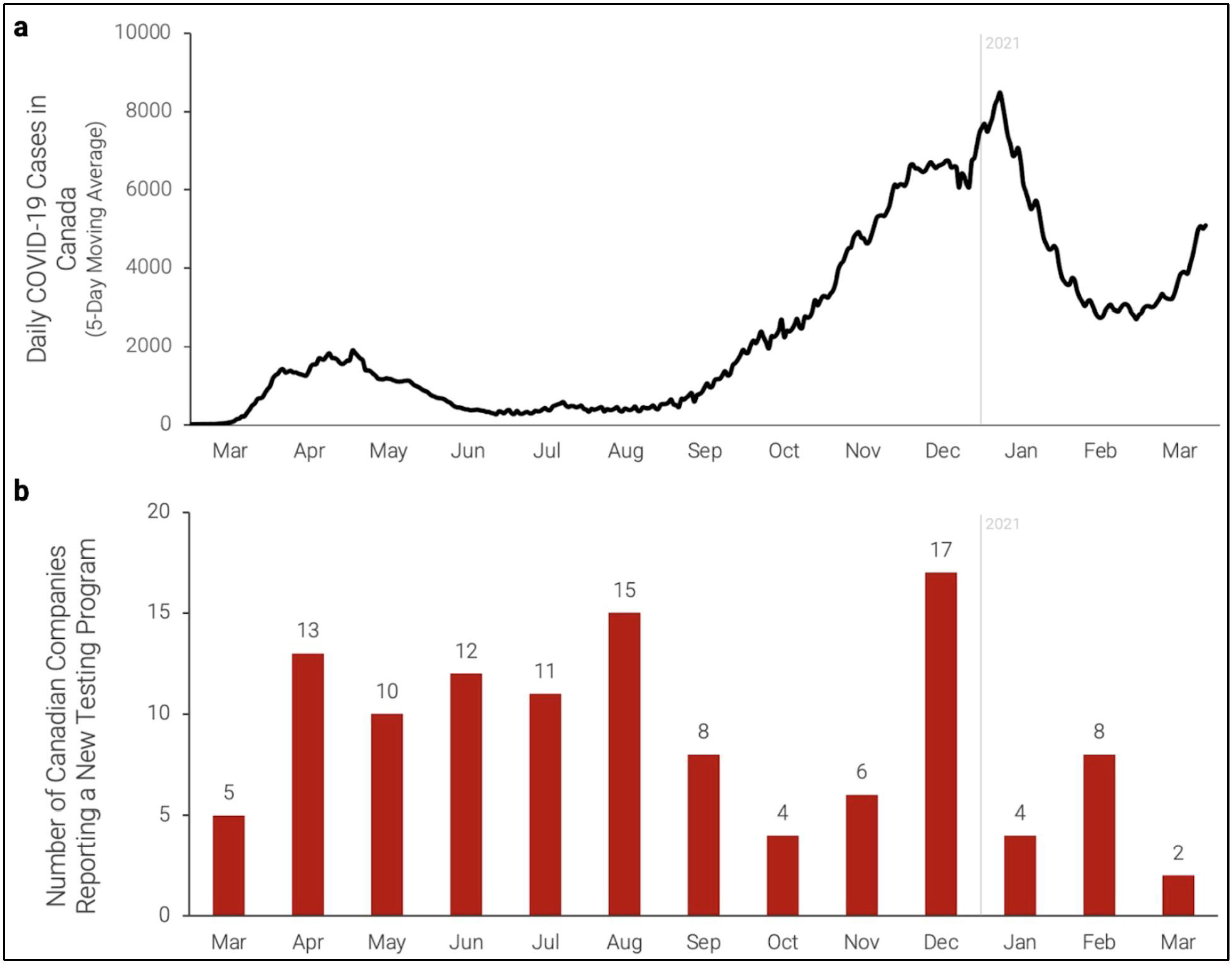
(a) Daily COVID-19 cases in Canada between March 1, 2020 and March 31, 2021. (b) Number of Canadian employers, among those we tracked, reporting a new workplace testing program each month between March 2020 and March 2021.

There were substantial differences in reported testing programs between sectors. Retail and customer-facing settings, indoor and mixed blue collar workplaces, and office settings had among the lowest proportions of employers reporting testing – fewer than 20% in each of these settings. A comparatively greater proportion of healthcare settings and universities (over 40% both within Canada and internationally) reported testing. These patterns were similar between Canadian and global employers.

## DISCUSSION

We used publicly-available data to monitor 2,240 employers for evidence of workplace testing. The volume of reported testing programs has increased since March 1, 2020, reflecting increased test availability and an improved understanding of how tests should be used. Even still, only 9.5% (n=110) of Canadian employers and 24.6% (n=266) of international employers we tracked reported testing as of March 31, 2021, highlighting opportunities to further improve workplace safety as businesses re-open.

In Canada, most testing programs (64.3%; n=74) were initiated in response to the first pandemic wave, but these responses were drawn out months after case volumes subsided. In contrast, the second “wave” of testing adoption that paralleled the second pandemic wave had a much faster ramp up, likely reflecting an improved understanding of SARS-CoV-2 epidemiology, and increased availability of tests and guidance for interested employers.

Antibody tests were more frequently used among employers earlier in the pandemic, as suggested by the range of dates on which these tests were adopted. However, at the time, there were misconceptions outside the scientific community around what these tests measured^13^ and whether they could be used as “immunity passports”^14^. Today, it is clear that antibody tests have substantial utility in seroprevalence studies (which map the extent of infection and vaccination on a population level^15^), but are not appropriate tools for identifying actively infectious employees^16^. Asking employees about vaccination status and diagnosis history, where legal, may be a more feasible strategy to gauge individual-level immunity.

Symptom-based screening measures will miss presymptomatic and asymptomatic infections which are key drivers of viral spread^17^, highlighting the importance of diagnostic testing in workplace safety controls. Nucleic acid tests – most often, lab-based PCR tests for diagnostic testing of individuals with suspected infection^18^ – were deployed in most (69.6%; n=174) testing programs that specified test type. However, high costs and slow turnaround times make frequent PCR testing of all employees impractical and less effective for flagging infectious employees in time to prevent spread^19^. Instead, after early skepticism around their utility, rapid antigen tests emerged during the second wave as preferable tests for screening asymptomatic employees^20,21^.

Public health leaders have advocated that employers conducting testing use a combined approach which includes both routine asymptomatic testing and confirmatory diagnostic testing^18^. However, we found that comparatively few employers specifying testing modality (9.2%; n=23) reported this combination, though some employers may be referring employees with positive antigen tests to public health units for diagnostic testing. Robust testing programs will be important in workplaces where vaccine uptake is low, especially in occupations that may have relatively low levels of vaccine confidence^7^. To maximize effectiveness and uptake, these testing programs should be paired with supporting measures including contact tracing systems, isolation and quarantine support, and paid sick leave^20 22^.

The rate of reported testing programs is concerningly low in retail and customer-facing settings (e.g., grocery stores), indoor blue collar work environments (e.g., factories, manufacturing plants, warehouses), and mixed blue collar work environments (e.g., mining sites, oil refineries, construction sites). Many of these settings have been associated with high risk of transmission and/or mortality^1,23,24^. Further, while effective SARS-CoV-2 vaccines are available and may reduce the need for ongoing workplace testing, low vaccine confidence is an issue among a number of occupations related to these settings^7^. Without robust testing programs, gaps in vaccination coverage will result in workforces that remain susceptible to outbreaks.

Employers with retail or customer-facing settings, or with blue collar work environments, can learn from the adoption of testing among food processing plants, which were associated with large outbreaks earlier in the pandemic^1^. Media and legal attention, and the fact that these plants are core nodes in essential supply chains, prompted a number of employers to deploy and report the use testing (see Figure 2)^25,26^. These testing programs may help compensate for any gaps in vaccine coverage resulting from hesitancy among food processing workers^7^.

**Figure 2.**
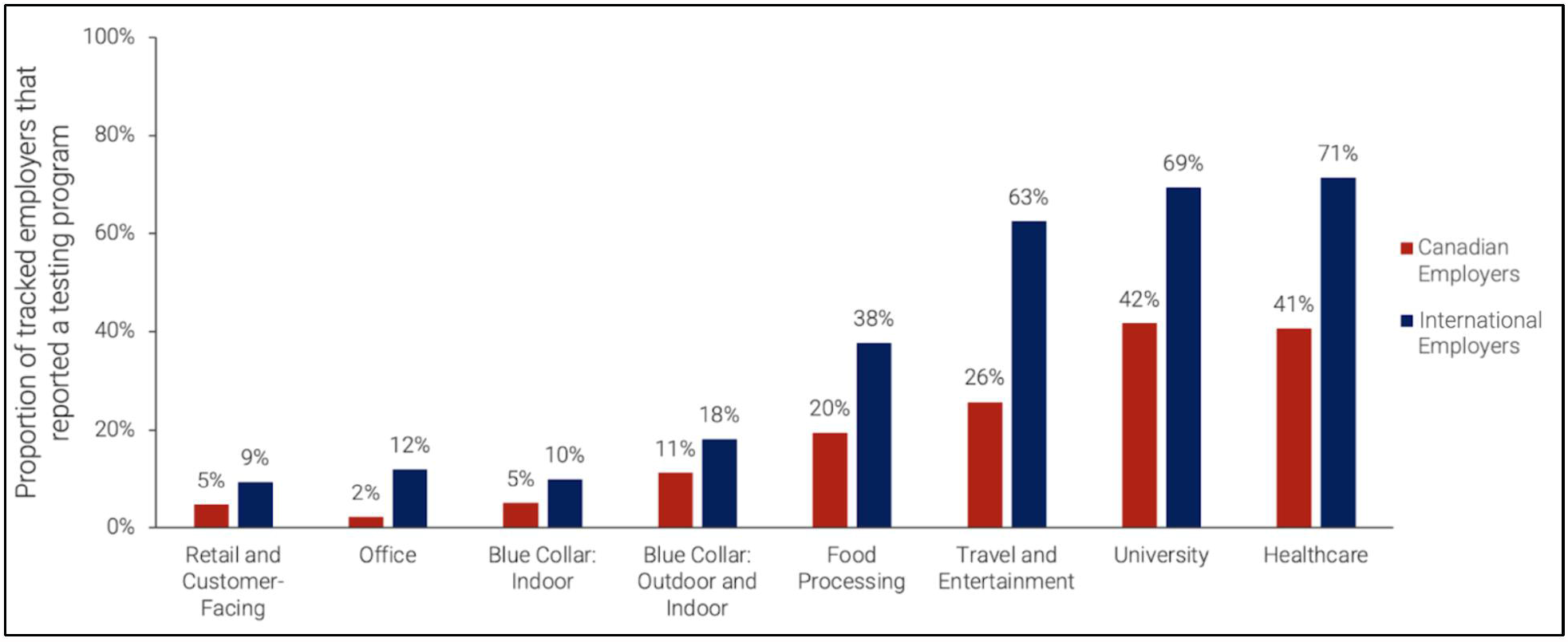
Proportion of tracked Canadian and international employers that had reported a workplace testing program as of March 31, 2021, stratified by type of workplace.

Using publicly-available data has unique advantages in tracking workplace testing programs. First, this approach is cheap, easily implementable, and can be scaled to thousands of employers. Second, our scraping process can be implemented in real-time, whereas employer surveys can take months to update^6^, helping public health officials and occupational health experts provide timely guidance. Finally, the use of public data means that there are no barriers to sharing detailed data broken down by sector, timing, or even individual employer, which would not be feasible with survey data collected with a promise of anonymity.

Nevertheless, our study has some limitations. First, we aimed to track large employers and our findings should not be generalized to small businesses. Small businesses comprise a large portion of most economies^27,28^ and can include occupations associated with particularly high risk of transmission (e.g., taxi drivers)^29,23^. Alternative strategies are required to understand precautions in these workplaces. Second, we likely underestimate the proportion of large employers testing employees because not all employers have an incentive to publicly disclose their efforts. Our findings therefore represent a lower bound of the proportion of employers conducting testing while surveys, which are likely skewed by non-response bias, represent an upper bound^6^.

Our findings highlight opportunities to further implement testing programs in high-risk workplaces and thereby improve workplace safety. In some cases, workforces in these high-risk settings may have low vaccine coverage due to low levels of vaccine confidence, underscoring the importance of including testing among other safety measures. In addition to promoting vaccine uptake, public health officials and occupational health experts should continue to support the adoption of testing in high-risk work environments, guided by a combination of publicly-available data and employer surveys.

## Data Availability

The data that support the findings of this study are available from the corresponding author, ND, upon reasonable request.

## Acknowledgements

We would like to acknowledge Madeleine Rocco, Austin Atmaja, and Abel Joseph for their role in data collection and screening, Tingting Yan for her role in funding acquisition, and Timothy Grant Evans for his role in motivating the project.

## REFERENCES

1. Murti M, Achonu C, Smith BT, et al. COVID-19 workplace outbreaks by industry sector and their associated household transmission, Ontario, Canada, January – June, 2020. J. Occup. Environ. Med. Published Online April 3, 2021.

2. Bank Of Canada. Business outlook survey—Spring 2020 [Bank of Canada website]. April 6, 2020. Available at: https://www.bankofcanada.ca/2020/04/business-outlook-survey-spring-2020/#Overview. Accessed October 20, 2020.

3. Poole SF, Gronsbell J, Winter D, et al. A holistic approach for suppression of COVID-19 spread in workplaces and universities. medRxiv 2020.

4. Campbell JR, Dion C, Uppal A, Yansouni CP, Menzies D. Systematic testing for SARS-CoV-2 infection among essential workers in Montréal, Canada: a prospective observational and cost assessment study. medRxiv 2021.

5. McKay SL, Tobolowsky FA, Moritz, ED, et al. Performance evaluation of serial SARS-CoV-2 rapid antigen testing during a nursing home outbreak. Ann Intern Med. Published Online 2021.

6. Wade NL, Aspinall MG. Back to the workplace: are we there yet? [The Rockefeller Foundation website]. April 2021. Available at: https://www.rockefellerfoundation.org/report/key-insights-from-employers-one-year-into-the-pandemic/. Accessed June 1, 2021.

7. King WC, Rubinstein M, Reinhart A, Mejia RJ. COVID-19 vaccine hesitancy January-March 2021 among 18-64 year old US adults by employment and occupation. medRxiv 2021.

8. Geers D, Shamier MC, Bogers S, et al. SARS-CoV-2 variants of concern partially escape humoral but not T-cell responses in COVID-19 convalescent donors and vaccinees. Sci. Immunol. 2021;6:59.

9. DeVito NJ, Richards GC, Inglesby P. How we learnt to stop worrying and love web scraping. Nature 2020;585:621–622.

10. Greenhalgh T, Jimenez JL, Prather KA, Tufekci Z, Fisman D. Schlooney R. Ten scientific reasons in support of airborne transmission of SARS-CoV-2. Lancet 2021;397:1603–1605.

11. Viera AJ, Garrett JM, Understanding interobserver agreement: the kappa statistic. Fam Med. 2005;37(5):360–363.

12. Google. How Search algorithms work [Google web site]. Available at: https://www.google.com/search/howsearchworks/algorithms/. Accessed May 20, 2021.

13. Lehmann, C. Viral, antibody test number policy sows confusion [WebMD website]. June 1, 2020. Available at: https://www.webmd.com/lung/news/20200601/viral-antibody-test-number-policy-sows-confusion. Accessed July 8, 2021.

14. Bramstedt KA. Antibodies as currency: COVID-19’s golden passport. J Bioeth Inq. 2020;17:687–689.

15. Bobrovitz N, Arora RK, Cao C, et al. Global seroprevalence of SARS-CoV-2 antibodies: a systematic review and meta-analysis. PLoS One 2021. In press.

16. Motley MP, Bennett-Guerrero E, Fries BC, Spitzer ED. Review of viral testing (polymerase chain reaction) and antibody/serology testing for Severe Acute Respiratory Syndrome-Coronavirus-2 for the intensivist. Crit. care explor. 2020;2(6):e0154.

17. Moghadas SM, Fitzpatrick MC, Sah P, et al. The implications of silent transmission for the control of COVID-19 outbreaks. Proc. Natl. Acad. Sci. U.S.A. 2020;117(30):17513–17515.

18. Government of Canada. Priority strategies to optimize testing and screening for COVID-19 in Canada: Report [Government of Canada website]. January 2021. Available at: https://www.canada.ca/en/health-canada/services/drugs-health-products/covid19-industry/medical-devices/testing-screening-advisory-panel/reports-summaries/priority-strategies.html. Accessed May 15, 2021.

19. Larremore DB, Wilder B, Lester E, et al. Test sensitivity is secondary to frequency and turnaround time for COVID-19 screening. Sci. Adv. 2021;7(1):eabd5393.

20. Schwartz KL, McGeer AJ, Bogoch II. Rapid antigen screening of asymptomatic people as a public health tool to combat COVID-19. CMAJ 2021;193(13):E449–E452.

21. Mina MJ, Andersen KG. COVID-19 testing: One size does not fit all. Science 2021;371:126–127.

22. Centers for Disease Control and Prevention. Contact tracing for COVID-19 [Centers for Disease Control and Prevention website]. February 25, 2021. Available at: https://www.cdc.gov/coronavirus/2019-ncov/php/contact-tracing/contact-tracing-plan/contact-tracing.html. Accessed March 14, 2021.

23. Magnusson K, Nygård K, Methi F, Vold L, Telle K. Occupational risk of COVID-19 in the 1st vs 2nd wave of infection. medRxiv 2020.

24. Chen Y, Glymour M, Riley A, et al. Excess mortality associated with the COVID-19 pandemic among Californians 18–65 years of age, by occupational sector and occupation: March through November 2020. PLoS One 2021;16(6):e0252454.

25. Fortin, J. After meat workers die of Covid-19, families fight for compensation [New York Times web site]. October 6, 2020. Available at: https://www.nytimes.com/2020/10/06/business/coronavirus-meatpacking-plants-compensation.html. Accessed December 27, 2020.

26. Taylor CA, Boulos C, Almond D. Livestock plants and COVID-19 transmission. Proc. Natl. Acad. Sci. U.S.A. 2020;117(50):31706–31715.

27. Government of Canada. Key small business statistics - Number 2019 [Government of Canada website]. December 10, 2019. Available at: https://www.ic.gc.ca/eic/site/061.nsf/eng/h_03114.html#figure12. Accessed June 1, 2021.

28. Arnold C. The foundation for economies worldwide is small business [International Federation of Accountants web site]. June 26, 2019. Available at: https://www.ifac.org/knowledge-gateway/contributing-global-economy/discussion/foundation-economies-worldwide-small-business-0. Accessed June 1, 2021.

29. McGran K. Amid COVID-19, Toronto taxi and Uber drivers say business is way down — and the risks are up [Toronto Star web site]. March 25, 2020. Available at: https://www.thestar.com/news/gta/2020/03/25/amid-covid-19-toronto-taxi-and-uber-drivers-say-business-is-way-down-and-the-risks-are-up.html. Accessed June 1, 2021.

