## Supplementary Data for "Uptake of SARS-CoV-2 workplace testing programs, March 2020 to March 2021"

Supplementary Table 1. Internally-developed list of workplace types.

| **Name** | **Description** | **Examples of Related NAICS Groups** |
| --- | --- | --- |
| Blue Collar: Indoor | Work environments in which skilled labour occurs indoors (distancing is difficult, increased transmission due to close-range droplets and long-range aerosols). | Factory/Manufacturing (31-33)<br>Warehouse (493)<br>Transportation/Supply Chain (48) |
| Blue Collar: Outdoor and Indoor | Work environments in which skilled labour occurs both indoors (distancing is difficult, increased transmission due to close-range droplets and long-range aerosols) and outdoors (distancing is more feasible, reduced transmission). | Utilities (22)<br>Oil and Gas (2111)<br>Mining (21)<br>Construction (23) |
| Food Processing | Food processing plants, agricultural facilities, and other such settings. Can contain a high number of people working within close proximity, in colder and drier air conditions. | Farms (11)<br>Food Manufacturing (311) |
| Healthcare | Workplaces that include heavy interaction between healthcare professionals and patients. | Healthcare and Social Assistance (62) |
| Retail and Customer-Facing | Workplaces where employees frequently interact with a large number of typically local customers or clients. | Retail Trade (44-45) |
| University | Workplaces where a large student body frequently interacts with each other and faculty. | Universities (611310) |
| Travel and Entertainment | Workplaces where a large number of customers from outside the locality heavily interact with employees. | Airlines (481111)<br>Airports (48811)<br>Traveler Accommodation (7211)<br>Casinos (71321) |
| Offices | Workplaces where employees work indoors with isolated interaction with other employees. Distancing is possible. | Finance and Insurance (52)<br>Scientific Research (5417)<br>Office Administrative Services (5611) |

Supplementary Table 2. Breakdown of tracked companies by workplace type and country.

| *Countries* | *Blue Collar: Indoor* | *Blue Collar: Outdoor and Indoor* | *Food Processing* | *Healthcare* | *Office* | *Retail and Customer-Facing* | *Travel and Entertainment* | *University* | *Other* | *Total* |
|---|---|---|---|---|---|---|---|---|---|---|
| *Canada* | 155 | 406 | 46 | 33 | 372 | 84 | 35 | 12 | 16 | 1159 |
| *United States* | 122 | 89 | 26 | 35 | 143 | 51 | 40 | 39 | 25 | 570 |
| *China* | 21 | 10 | 10 | 1 | 42 | 3 | 4 | 0 | 0 | 91 |
| *Brazil* | 11 | 14 | 5 | 1 | 13 | 10 | 3 | 0 | 0 | 57 |
| *Japan* | 25 | 7 | 0 | 0 | 11 | 2 | 0 | 1 | 0 | 46 |
| *India* | 9 | 8 | 0 | 1 | 6 | 3 | 1 | 3 | 3 | 34 |
| *Germany* | 11 | 3 | 1 | 1 | 9 | 3 | 1 | 1 | 0 | 30 |
| *United Kingdom* | 5 | 4 | 5 | 2 | 6 | 2 | 2 | 1 | 1 | 28 |
| *South Korea* | 8 | 4 | 0 | 0 | 7 | 0 | 0 | 0 | 2 | 21 |
| *Switzerland* | 6 | 2 | 2 | 0 | 6 | 4 | 0 | 0 | 0 | 20 |
| *France* | 2 | 4 | 2 | 0 | 4 | 4 | 1 | 1 | 1 | 19 |
| *Spain* | 1 | 7 | 0 | 1 | 4 | 1 | 3 | 2 | 0 | 19 |
| *Mexico* | 1 | 3 | 3 | 0 | 3 | 4 | 2 | 0 | 1 | 17 |
| *Netherlands* | 4 | 1 | 1 | 0 | 5 | 2 | 0 | 1 | 2 | 16 |
| *Italy* | 3 | 4 | 0 | 0 | 4 | 1 | 1 | 1 | 1 | 15 |
| *Australia* | 2 | 4 | 0 | 0 | 4 | 4 | 0 | 0 | 1 | 15 |
| *Russia* | 1 | 9 | 0 | 0 | 1 | 0 | 1 | 1 | 0 | 13 |
| *Hong Kong* | 1 | 3 | 0 | 0 | 6 | 0 | 1 | 0 | 2 | 13 |
| *Saudi Arabia* | 1 | 3 | 1 | 0 | 3 | 0 | 0 | 0 | 0 | 8 |
| *Belgium* | 1 | 1 | 1 | 0 | 2 | 0 | 0 | 1 | 0 | 6 |
| *Other* | 3 | 12 | 4 | 0 | 5 | 2 | 4 | 10 | 3 | 43 |
| *Total* | 393 | 598 | 107 | 75 | 656 | 180 | 99 | 74 | 58 | 2240 |
